# Blood clot resolution using active fluid exchange in the treatment of intraventricular hemorrhage

**DOI:** 10.1101/2023.07.04.23292127

**Authors:** Mette Haldrup, Anders Mølgaard Jakobsen, Mads Rasmussen, Stig Dyrskog, Claus Ziegler Simonsen, Mads Grønhøj, Frantz Rom Poulsen, Anders Rosendal Korshøj

## Abstract

**Objective:** Intraventricular hemorrhage (IVH) is a severe condition with poor outcome and high mortality. New treatments are warranted to facilitate clot removal and accelerate recovery of IVH patients. We examined the effect to intraventricular lavage on IVH resolution, clearance and kinectics. The study is a post-hoc study on a recently performed randomized study, where intraventricular lavage using controlled irrigation and aspiration was tested against passive drainage. The present study describe post hoc analysis of the Active Study to determine the effect of intraventricular lavage on IVH resolution clearance and kinetics.

**Method:** postPost-hoc analysis using data from the multi-center, randomized, controlled clinical safety/feasibility trial (Active Study, NCT05204849) including 21 patients with IVH. IVH clearance was assessed using volume segmentation of serial CT scans. Clot resolution kinetics were determined and correlated with irrigation rates.

**Results:** The data set consisted of 77 sequential CT scans from 18 patients (3 of the 21 patients included in the Active study were excluded due to rebleeding or only one CT scan performed). Baseline characteristics were equal in the two groups. Clot resolution followed 0-order kinetics. The clot half-life was 3.9 days in the group treated with intraventricular lavage compared to 5.3 days in the control group treated with passive drainage (p = 0.6). We did not observe accelerated clot resolution in the ventricles containing the catheter tip (3.9 days vs 3.6 days, p = 0.9). In the intervention group, clot resolution was increased by 0.05% per ml increase in irrigation (slope 1.576 ml/cm3 (95%CI 0.55-2.6) p=0.03)

**Conclusion:** We found that IVH clot resolution followed 0-order kinetics. No significant differences in clot half-life were observed between the two groups. However, the clot resolution rate was positively correlated with the irrigation rates.

## Introduction

Intraventricular hemorrhage (IVH) is a life-threatening condition with poor neurological outcomes and a high mortality rate^1,2^. Blood in the ventricles contribute to morbidity in different ways: 1) acute hydrocephalus due to blockage of cerebrospinal fluid (CSF) flow, 2) chronic hydrocephalus due to blood degradation products embedded in the arachnoid granulations, and 3) cognitive dysfunction due to the inflammatory reaction from blood breakdown products^3,4^. Currently, the treatment of IVH relies on supportive care with CSF drainage to decrease intracranial pressure (ICP) and to passively facilitate hematoma evacuation using an external ventricular drain (EVD)^3^.

Recent studies have shown that intraventricular fibrinolysis (IVF) can accelerate clot removal, leading to improved survival and functional outcomes^2,5^. No significant treatment advances have been made in the past decades. Minimally invasive techniques clot removal could mitigate better outcome are lacking.

A new technology, IRRAflow®, has been introduced to accelerate IVH clearance and recovery of IVH patients through active and controlled perfusion of the ventricular system using a dual lumen catheter. IRRAflow® performs periodic, controlled irrigation and aspiration with physiological saline, while simultaneously monitoring the ICP (Fig. 1). The safety and efficacy of IRRAflow® was recently evaluated in a randomized controlled trial against a standard external ventricular drain (EVD) in IVH patients. The study aimed to evaluate the efficacy of intraventricular lavage for faster clearance of blood. However, the study was terminated early due to a significantly increased number of severe adverse events (SAEs) in the intervention group (Incidence rate ratio = 6.0, 95% CI 1.38-26.11, p=0.012). Although concerns are associated with the current version of IRRAflow®, it is plausible that the concept of active ventricular lavage holds significant potential. Intraventricular lavage has been tested in different setups, especially in patients suffering subarachnoid hemorrhage (SAH). The studies report that intraventricular lavage was safe to use and reduced the risk of vasospasms using cisternal irrigation strategies^6^.

**Figure 1.**
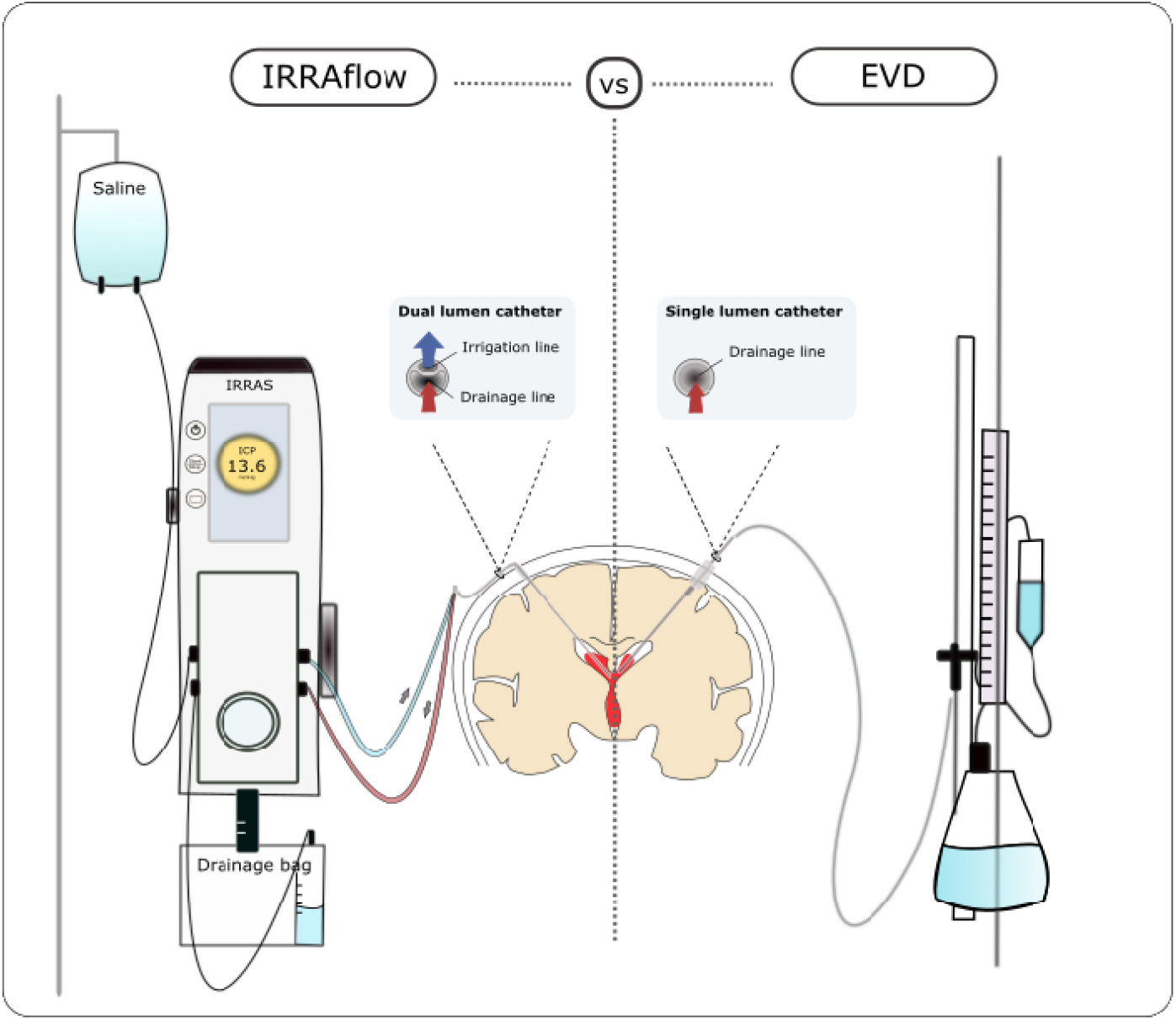
The IRRAflow® device showing the control unit, the tube system, the dual lumen catheter and the collection bag system (left), and the external ventricular drain (EVD) with passive gravity driven drainage through a single lumen catheter (right).

In this paper, we aim to explore the hypothesis of increased clot resolution following intraventricular lavage by conducting a thorough assessment of IRRAflow®’s ability to clear IVH effectively. We present a post hoc volumetric evaluation of IVH wash-out with active irrigation and aspiration compared to standard EVD with gravity-driven CSF drainage, and we correlate the amount of irrigation used with the rate of blood clearance.

## Methods

### Study design and patient population

We conducted a post-hoc analysis using data from a multi-center, randomized, controlled clinical safety/feasibility trial (Active Study, NCT05204849) performed at Aarhus University Hospital and Odense University Hospital in Denmark. The study aimed to test the primary hypothesis that continuous ventricular perfusion with IRRAflow® would lead to a lower incidence of catheter occlusions compared to standard EVD treatment in patients with IVH. Secondary outcomes included adverse events and IVH clearance rate. Patients aged ≥18 years with a primary or secondary IVH and indication for CSF drainage were assessed for eligibility. Inclusion criteria required an IVH Graeb score > 3 and a head CT within 24 hours before inclusion. Pregnant or nursing women and patients with fixed and dilated pupils were excluded. The study was designed to enroll 58 patients but was terminated early due to safety concerns after enrolling 21 patients (11 in the intervention group and 10 in the control group). This post hoc analysis is based on volumetric assessments of the 21 enrolled patients.

### Volumetric assessment and image processing

Volumetric assessments were performed using head computed tomography (CT) scans with slice thickness ranging from 0.625 mm to 1 mm. Each patient underwent a CT scan prior to catheter placement and was subsequently scanned 3 to 5 times during the initial 9 days of treatment (Table 1). All exams were saved in standard Digital Imaging and Communications in Medicine (DICOM) format and exported from Enterprise Imaging (AGFA HealthCare, Belgium).

**Table 1.** Baseline characteristics

|  | Intervention | Control | p-value |
| --- | --- | --- | --- |
| Age, mean (SD) | 66 (14.4) | 64 (11.4) | 0.722 |
| Male (n) | 7 | 5 | 0.620 |
| Female (n) | 2 | 4 |  |
| IVH type |  |  |  |
| SAH with intraventricular extension (n) | 4 | 3 | 0.637 |
| ICH with intraventricular extension (n) | 4 | 6 |  |
| Primary IVH (n) | 1 | 0 |  |
| Graeb score, mean (SD) | 8.6 (1.9) | 9.1 (1.7) | 0.558 |
| IVH volume ml (baseline CT), median (range) | 27 (3-64) | 14 (8-103) | 0.759 |

The CT scans were segmented by a resident neurosurgeon (MH) and a medical engineer specialized in medical 3D visualization and modulation (AMJ) using Mimics Medical 25.0 software (Materialize, Belgium). A Hounsfield Unit (HU) threshold of 60-100 was applied to semi-automate the segmentation process and a margin of 10 HU above and below the standard 70-90 HU was used due to the variable HU thresholds within the clot (5, 6).

The CT scan segmentation was performed in four steps: (i) an automatic creation of a 3D mask within the 60-100 HU threshold, (ii) manual isolation of the region of interest, (iii) semi-automated edit of the region of interest with a spherical radius tool to finalize the 3D mask and correct possible false positive areas or missing clot areas, and (iv) transformation of the 3D mask into a 3D object from which the volumes were calculated (Fig. 2).

**Figure 2.**
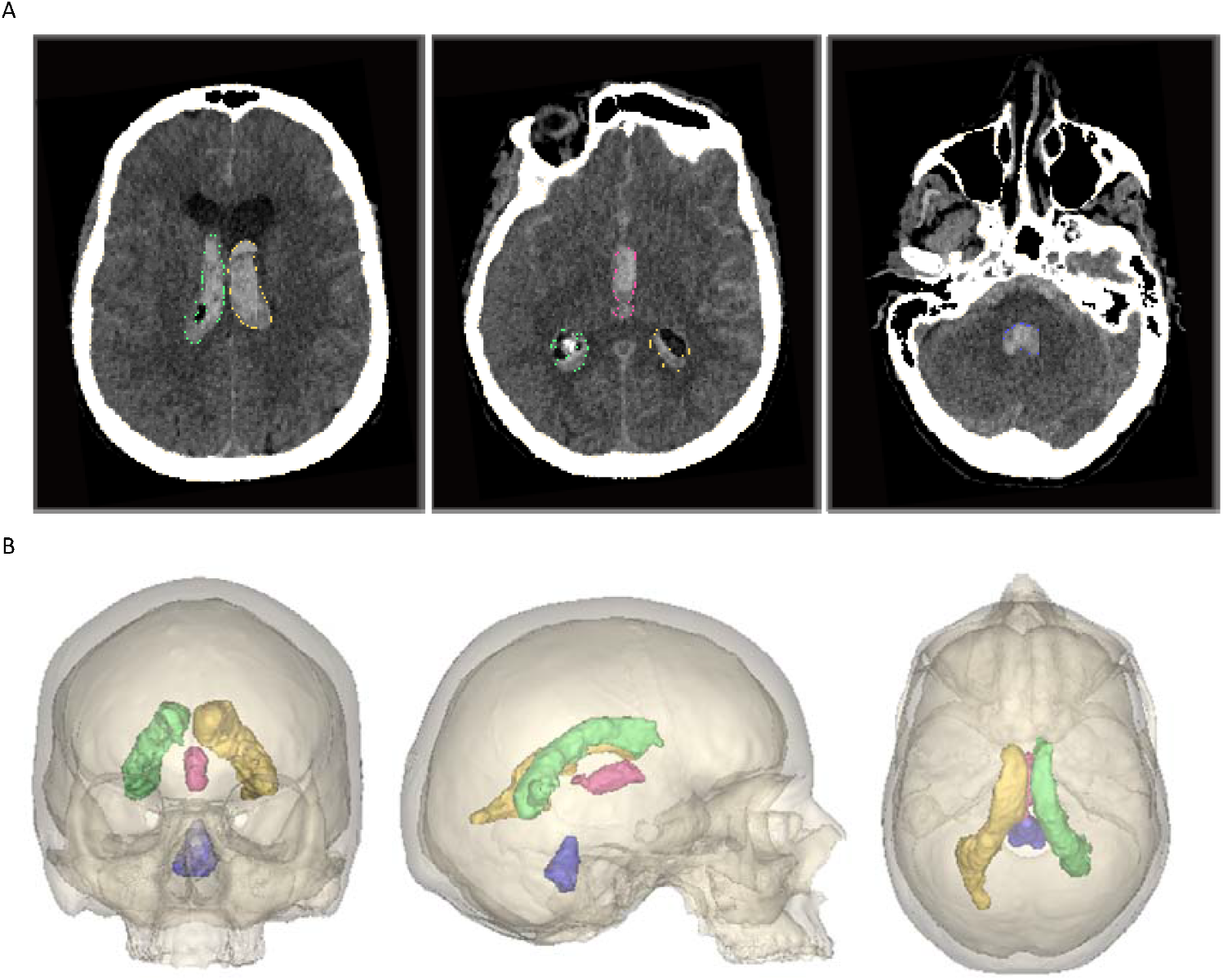
Segmentation results of the semiautomated 3D parts. A Axial views showing the lateral ventricles, third ventricle and lateral ventricles and fourth ventricle (green = blood clot right lat. ventricle, yellow = blood clot left lat. ventricle, pink = blood clot third ventricle, and purple = blood clot fourth ventricle) and B a 3D reconstruction of the ventricular blood

IVH volumes were calculated in three ways: (1) total amount of blood within the ventricular system, (2) amount of blood within the ventricle containing the tip of the irrigation catheter to evaluate the potential effect of the irrigation, and (3) volume of diluted blood sediment (Table 1). The IVH volumes were compared between the intervention and control groups. Additionally, we correlated the irrigation rates with the rate of clot removal for the intervention group between each scan for the first 9 days of treatment.

### Statistical analysis

To evaluate the kinetics of intraventricular clot resolution, we used a linear mixed model and random coefficient model to compile serial group representation of clot volume over time. Time zero (t0) was defined as the time of the baseline head CT scan prior to catheter placement. We calculated the time (t) of each subsequent head CT from the baseline scan and rounded it to the nearest hour. Further assessment of the kinetic of clot resolution was assessed using the local estimated scatterplot smoothing (LOESS) curve.

We determined the absolute rate of clot resolution by calculating the change in clot volume (ml) at time t compared to the baseline volume at t0. We assumed a constant absolute volume resolution over time and calculated the clot half-life as the time required to achieve a 50% volume reduction relative to the initial CT. The clot half-life was calculated for each group, and we used post-hoc commands to calculate 95% CI.

To evaluate the potential difference between the intervention group and the control group, we compared the slopes of the graph in each group. We also calculated the total volume of blood sediment in the ventricular system as the absolute increase in volume per day (ml). To obtain a more precise and localized evaluation of the wash-out potential of the irrigation system, we calculated the clot half-life for the specific ventricle containing the catheter tip.

For patients treated with IRRAflow®, we correlated clot resolution to the amount of irrigation saline per day using a mixed effect model (fixed effect model for amount of irrigation and a random effect model for patient id). In this analysis, we correlated the amount of irrigation between every scan to the wash-out of blood during the first nine days of treatment. The irrigation rate was as standard most of set at 180 ml/hour. Only in case of marginal ICP control the irrigation rate was lowered. The irrigation and drainage were only active when the intracranial pressure (ICP) was above the user-defined threshold (“Drain above”). Therefore, we calculated the actual amount of irrigation per day and used it in the correlation between the amount of irrigation and clot resolution. These data were analyzed using a mixed model with irrigation volume and duration as fixed effects and patient id as a random effect with an exponential residual variance structure varying over days after t0.

## Results

The data set consisted of 77 sequential CT scans of 18 patients. Two patients of the original study cohort of 21 patients were excluded from the analysis as they only had one CT scan performed before they died, and 1 patient was excluded due to two severe rebleeding episodes at days 1 and 3 (Supplementary 1). The baseline characteristics were equal between the two groups. Baseline characteristics of the included patients are shown in Table 1. Clot resolution kinetics were found to be constant over time and followed 0-order kinetics which was confirmed by the LOESS curve.

The median (interquartile range [IQR]) clot volumes were 27 ml (3-65 ml) in the intervention group and 15 ml (8-103 ml) in the control group. In 2 of 18 patients (11.1%) we observed a 4.4% and 16.2% clot volume expansion, respectively, during the first two days with no sign of rebleeding.

The mean and standard error (SE) clot half-life was 3.9 (1.36) days in the intervention group compared to 5.3 (2.82) days in the control group (p = 0.6) (Fig. 3a). Clot half-life in the ventricle containing the catheter tip was 3.9 days (SE 1.8 days) in the intervention group compared to 3.6 days (SE 3.1 days) in the control group (p = 0.9) (Fig. 3b). The mean daily irrigation volume was 1094 ml/day (range 0-4166 ml/day) in the intervention group. Clot resolution was increased by 0.05% per ml increase in irrigation (slope 1.576 ml irrigation/ml clot removal (95%CI: 0.55-2.6, p=0.03) illustrating that when observing two participants with one unit difference in irrigation amount on the same day, an expect expected difference in clot removal would be 1.76 ml (Fig. 4).

**Figure 3.**
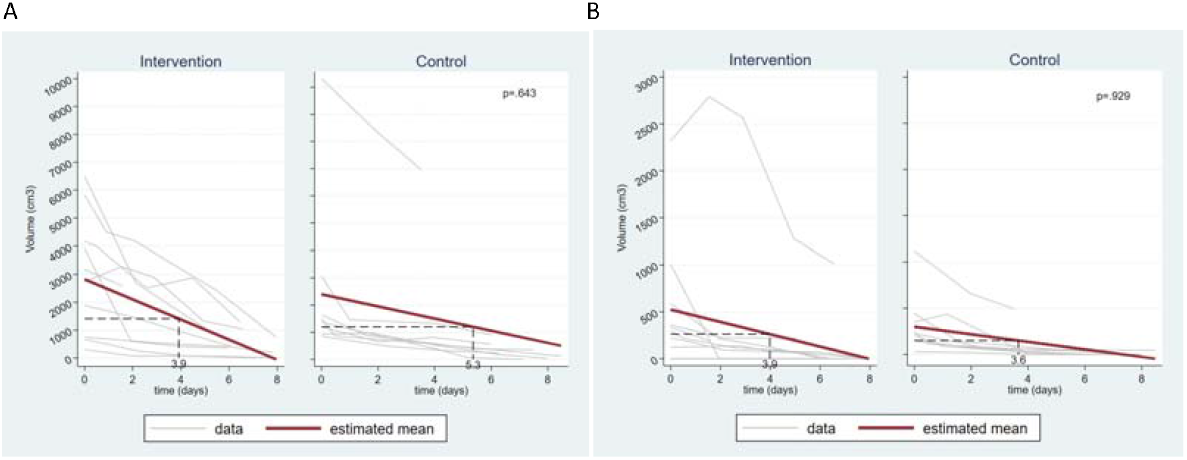
a. Absolute clot removal (mm3) of the total hematoma in the ventricular system over the fist 9 days after catheter placement. b. Absolute clot removal (mm3) of the ventricle containing the catheter tip over the fist 9 days after catheter placement. Gray lines: clot removal for each individual patient. The red line: Fitted accumulated line, all patients.

**Figure 4.**
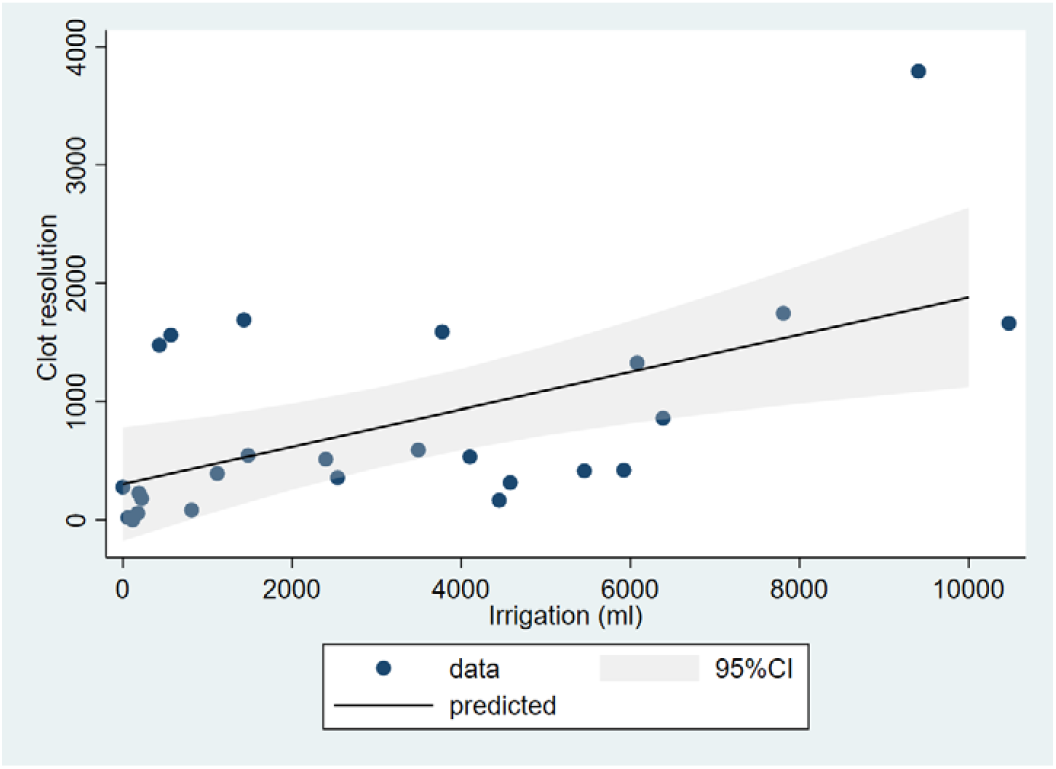
Correlation between amount of irrigation and clot wash out in only the IRRAflow® treated patients with significantly increased clot washout following the amount of irrigation (slope 1.576 ml/cm3 (95%CI 0.55-2.6) p=0.03)

The change in total sedimentation volume was not significantly different between the two groups (0.3 ml/day, SE 0.3 ml/day) in the intervention group vs 0.2 ml/day (SE 0.4 ml/day) in the control group (95%CI:-1.008-1.262, p = 0.8).

## Discussion

We did not find a statistically significant difference in the clot resolution rate between the irrigation group and the standard EVD group (3.9 days vs 5.3 days, p = 0.6). Neither did we observe a difference when comparing the ventricles containing the catheter tip where the irrigation was applied when comparing irrigation to the passive EVD (3.9 days vs 3.6 days, p = 0.9). We did observe a significant correlation between IVH clearance rate and the amount of irrigation, indicating that the mechanical effect of irrigation alone has a positive effect of clot removal. In the Active study the catheter was per protocol placed in the ventricle containing the least blood to reduce risk of occlusions and thereby not irrigating directly into the hematoma unless blood was present in both lateral ventricle and third ventricle.

It has been proposed that EVD treatment alone could slow the rate of clot resolution by removing the tissue plasminogen activator (tPA) liberated from the clot into the CSF^7^. The same could be suspected for irrigation with saline alone. Conversely, the injection of thrombolytic agents (ie. tPA) into the ventricular space would increase the rate of clot resolution and has been found to reduce time for clearance of ventricular blood from 8 to 4 days^2,8^. Use of fibrinolytic agent and lavage has further been tested in patients with SAH and found to significantly increase clot wash out (p<0.05), lower mortality rates (p<0.05), lower risk of cerebral infarction (p<0.05) and increase functional outcome after 6 month (p<0.05)^6^. Currently, different studies are evaluating the IRRAflow® system in combination with tPA treatment (ARCH (NCT05118997), AFFECT and DIVE trials). No results are published yet, but irrigation in combination with tPA could be a promising combination. Therefore, to improve the mechanical effect of clot wash out by the IRRAflow®, tissue plasminogen activator (tPA) can be administered to the irrigation saline and further by adding tPA, the catheter tip could potentially also be placed directly in the clot without risk of occlusions. However, ACTIVE study was evaluating safety and efficacy in the use of the IRRAflow® device and tPA was not applied.

IVH clot resolution was in our study found to follow a zero-order kinetic and was constant over time with a clot half-life proportional to the initial clot volume, which supports previous findings^7^. Previous studies also observed a latency period of 1-2 days in clot resolution. This was further investigated in a study from 2004 which concluded that this latency could be explained by the transformation of the precursor plasminogen to plasmin^9^. Plasmin is carried into the ventricles in its precursor form, plasminogen, as a normal constituent of blood and the amount of this enzyme is proportional to the clot volume. Plasminogen is then converted to the active form by tPA, which is released by the endothelium of vessels in the meninges and the ependyma as well as the leukocytes and platelets contained within the clot. This indicates that the amount of tPA is dependent on the severity of the hemorrhage^10^. In our study, we did not observe this latency period. However, the first CT scan after the baseline scan was performed 2 days after the baseline scan and thereby potentially the latency period could have been missed. Neither did we observe a latency period in the intervention group receiving intraventricular irrigation which in theory could wash out endogenous tPA.

In 11.1 % of the cases, we found the clot volume to expand with no sign of rebleeding within the first 2 days. This phenomenon has previously been described. They observed that close to 30 % of the study population demonstrated a 5% increase (range 6-15.8%) in clot volume beyond their initial clot volume^7^.

## Study limitations

Due to necessary early termination, the present study was underpowered for several endpoints including clot resolution kinetics. This represents a significant limitation in the ability to make firm conclusions. The small sample size also affected the ability of statistical analyses to assess the kinetics of the clot resolution. Former studies have found clot resolution to follow zero order kinetics and a LOESS curve assessment confirmed zero order kinetics in our dataset. Another limitation in this study was that the catheters were placed, if possible, outside of the clot to reduce risks of occlusions especially in the control group. The actual mechanical effect of clot wash out is therefore potentially higher than observed in this study. Furthermore, participants with foramen Monroe block were treated with standard EVD in the contralateral ventricle, regardless of randomization which could potentially have affected the clot resolution differently in the two groups.

## Conclusion

No significant differences in clot half-life were observed between the two groups. However, the rate of clot resolution was positively correlated with irrigation rates, and we cannot exclude that a adequately powered study would indicate a benefit of irrigation. Further studies should be conducted to elucidate the potential of ventricular lavage in combination with tPA.

## Data Availability

All data produced in the present work are contained in the manuscript

## Other information

### Registration

ClicalTrials.gov identifier: NCT05204849, registered 15 December 2021, updated 24 January 2022

### Protocol

The published full protocol can be found https://doi.org/10.1186/s13063-022-07043-9 or in Supplementary Material 1

## Supplementary material

**Supplementary material 1.**
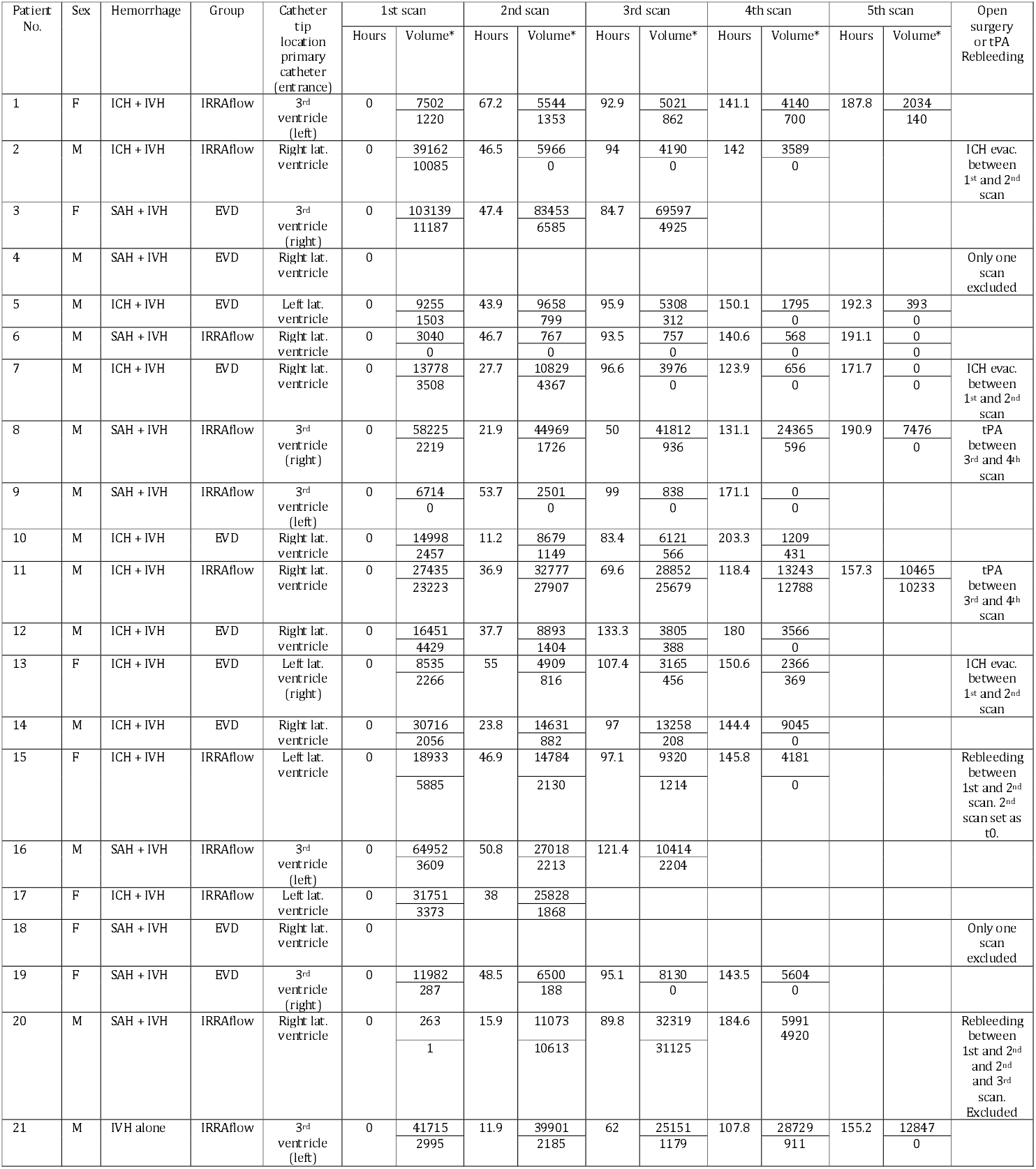
Baseline characteristics, time of computed tomography (CY) and clot volumes. EVD, external ventricular drainage; Volume, volume Mm^3^ ; ICH, Intracerebral hemorrhage; SAH, Subarachnoid hemorrhage.

